# Circulating Tau Profiles in Pediatric and Adult Patients with Spinal Muscular Atrophy

**DOI:** 10.64898/2025.12.19.25342711

**Authors:** Leillani L. Ha, Sunayana Mitra, Doreen T. Ho, Becky Fillingham, James D. Berry, Kathryn J. Swoboda, Christiano R. R. Alves

## Abstract

**Objective:** To determine alterations in circulating Tau and phosphorylated Tau (pTau) profiles in pediatric and adult patients with spinal muscular atrophy (SMA).

**Methods:** Circulating total Tau, pTau-181, pTau-217, pTau-262, and pTau-396 concentrations were measured across three cohorts: 1) adults including healthy controls, SMA patients, and ALS patients; 2) pediatric SMA patients and age-matched controls; and 3) pediatric SMA patients treated with onasemnogene abeparvovec.

**Results:** Distinct alterations in circulating Tau species were detected in adult SMA and ALS. Among all measurements, pTau-262 emerged as the only species specifically elevated in adult SMA, while total Tau levels were comparable between adult SMA and controls but significantly increased in ALS. Tau alterations were not consistently observed in pediatric SMA, although a small subset showed elevated levels, underscoring the value of individualized biomarker monitoring upon diagnosis. In gene-therapy–treated infants, Tau levels increased transiently several weeks after onasemnogene abeparvovec injection, paralleling previously described neurofilament kinetics and suggesting acute, treatment-associated neuronal stress.

**Conclusions:** Circulating Tau, particularly pTau-262, may serve as a disease-relevant biomarker in adult SMA, while pediatric profiles appear more heterogeneous. Transient Tau elevations after gene therapy may reflect acute neuronal vulnerability and warrant further investigation.

## Introduction

The development and application of reliable biomarkers have become increasingly critical in neurology as therapeutic interventions expand, particularly for neuromuscular disorders. Tau, a microtubule-associated protein essential for maintaining axonal structure and transport, has emerged as an informative marker of neuroaxonal injury^1^. Under normal physiological conditions, tau phosphorylation supports microtubule binding and cytoskeletal plasticity. However, dysregulated kinase activity in different diseases can lead to aberrant Tau phosphorylation, including the residues T181 (pTau-181), T217 (pTau-217), T262 (pTau-262), and S396 (pTau-396)^2,3^. These modifications can impair axonal transport, and promote the formation of neurofibrillary aggregates that contribute to neuronal dysfunction^2,3^. Elevated pTau species have been well documented in neurodegenerative diseases including amyotrophic lateral sclerosis (ALS), a severe the neuromuscular conditon^4–11^. However, a distinct non-aggregating tau-mediated toxicity at the neuromuscular junction pathology is expected based on pre-clinical studies for other neuromuscular conditions such as spinal muscular atrophy (SMA), a leading infantile genetic cause of death worldwide^12^. Our understanding of circulating Tau and pTau levels as potential accessible peripheral biomarkers of ongoing neuronal death in SMA is still very limited. Given its mechanistic role, Tau represents a compelling candidate for further study in this context but with limited clinical evidence. Thus, we aimed to determine alterations in circulating Tau and pTau profiles in cohorts of both pediatric and adult patients with SMA.

## Methods

### Study Approval and Subjects

Written informed consent or parental consent were obtained from all participants under Institutional Ethics Review Boards at the University of Utah (protocol 8751) and Massachusetts General Hospital (protocols 2025P001475, 2016P000469, 2021P000822, and 2006P000982). We queried our local databases^13,14^ for all serum and plasma samples available from SMA subjects above 14 years old (Cohort 1: adults) or under 2 years of age (Cohort 2: pediatric) and not receiving any SMN-targeted molecular or gene therapy at the time of sample collection. Moreover, we used a subset from a previously published study from our group including all samples available for pediatric SMA patients treated with gene therapy at our own site (Cohort 3)^14^. We included healthy control subjects for the natural history cohorts (Cohorts 1 and 2) as well as samples collected on site from ALS patients for the adult cohort (Cohort 1). **Table 1** presents the distribution of age, sex and SMN2 copies for these three independent cohorts.

**Table 1.** Demographic table of SMA, ALS, and control subjects in each cohort studied.

|  | Cohort 1 (n = 59) |  |  | Cohort 2 (n = 42) |  |  | Cohort 3 (n = 8) |  |
| --- | --- | --- | --- | --- | --- | --- | --- | --- |
|  | Control<br>(n = 17) | SMA<br>(n = 22) | ALS<br>(n = 20) | Control<br>(n = 10) | 2 SMN2<br>copies<br>(n = 22) | 3 SMN2<br>copies<br>(n = 10) | AVEXIS<br>(n = 4) | Co-Therapy<br>(n = 4) |
| **Gender** |  |  |  |  |  |  |  |  |
| Male | 7 (41.2%) | 11 (50%) | 12 (60%) | 7 (70%) | 12 (55%) | 7 (70%) | 2 (50%) | 2 (50%) |
| Female | 10 (58.8%) | 11 (50%) | 8 (40%) | 3 (30%) | 10 (45%) | 3 (30%) | 2 (50%) | 2 (50%) |
| **Age** | 45.18 ±<br>18.45 (y) | 35.1 ±<br>15.12 (y) | 61.85 ±<br>11.02 (y) | 88.1 ±<br>174.58 (d) | 224 ±<br>194.78 (d) | 416.2 ±<br>166.50 (d) | 217.56 ±<br>154.69 (d) | 120.05 ±<br>72.38 (d) |
| **Age range** | 24-74 | 14-65 | 35-76 | 0-554 | 0-780 | 125-692 | 24-556 | 9-252 |
| **SMN2 copies** |  |  |  |  |  |  |  |  |
| 1 | - | 2 (8%) | - | - | - | - | - | - |
| 2 | - | 1 (4%) | - | - | 22 (100%) | 0 (0%) | - | 4 (100%) |
| 3 | - | 9 (36%) | - | - | 0 (0%) | 10 (100%) | 2 (50%) | - |
| 4+ | - | 10 (45%) | - | - | - | - | 2 (50%) | - |

### Circulating Tau, pTau and neurofilament levels

Circulating total Tau, pTau-181, pTau-217, pTau-262, and pTau-396 concentrations were measured using commercially available and validated immunoassays. Different kits required different dilution factors. Total Tau levels were measured using Human Tau ELISA Kit (Catalog # NBP2-62749; Novus Biologicals LLC a Bio-Techne Brand, CO, USA) at the 1:4 dilution factor. For pTau-181, pTau-217 and pTau-262, a LEGENDplex™ Pre-defined Human Phospho-Tau Panel (Catalog # 741507; BioLegend; CA, USA) was performed at the 1:2 dilution factor. For pTau-396, a Human Tau (Phospho) [pS396] ELISA Kit (Catalog # EEL188; Invitrogen / Thermo Fisher Scientific; MA, USA) was performed at the 1:4 dilution factor. For Cohorts 1 and 2, all analysis were performed using serum samples. For Cohort 3, all analysis were performed using plasma samples due to limited number of available serum samples. Neurofilament levels analyzed in Cohort 3 are the same data previously reported in our previous studies using a pNF-H enzyme-linked lectin assay (ProteinSimple, CA, USA) and a single-molecule array (Simoa) NF-light Advantage Kit (Quanterix, Billerica, MA, US)^14^.

### Statistical Analysis

Data is presented as mean ± 95% confidence interval (CI) as well as individual participant data. Statistical comparisons were performed using one-way ANOVA followed by multiple comparisons by controlling false discovery rate using two-stage step-up method of Benjamini, Krieger and Yekutieli. Critical value was set as p < 0.05.

## Results

To assess whether circulating Tau levels are altered in adults with SMA, we analyzed serum samples from 17 healthy controls and 22 untreated adult SMA patients (natural history cohort). We also included 20 adults with ALS as a disease control group. Total Tau, pTau-181, pTau-217, pTau-262, and pTau-396 concentrations were quantified in all samples (**Figure 1A**). Total Tau levels were comparable between healthy controls and SMA patients, whereas ALS patients showed significantly elevated total Tau compared with both groups (**Figure 1B**). No significant differences were observed among the three groups for pTau-181 or pTau-217 (**Figure 1C–D**). Interestingly, SMA patients, but not ALS patients, displayed higher pTau-262 levels relative to healthy controls (**Figure 1E**). In contrast, ALS patients exhibited increased pTau-396 levels, a change not observed in SMA (**Figure 1F**). Together, these results highlight distinct alterations in circulating Tau species across SMA and ALS, with pTau-262 emerging as the only Tau measurement specifically elevated in adults with SMA.

**Figure 1.**
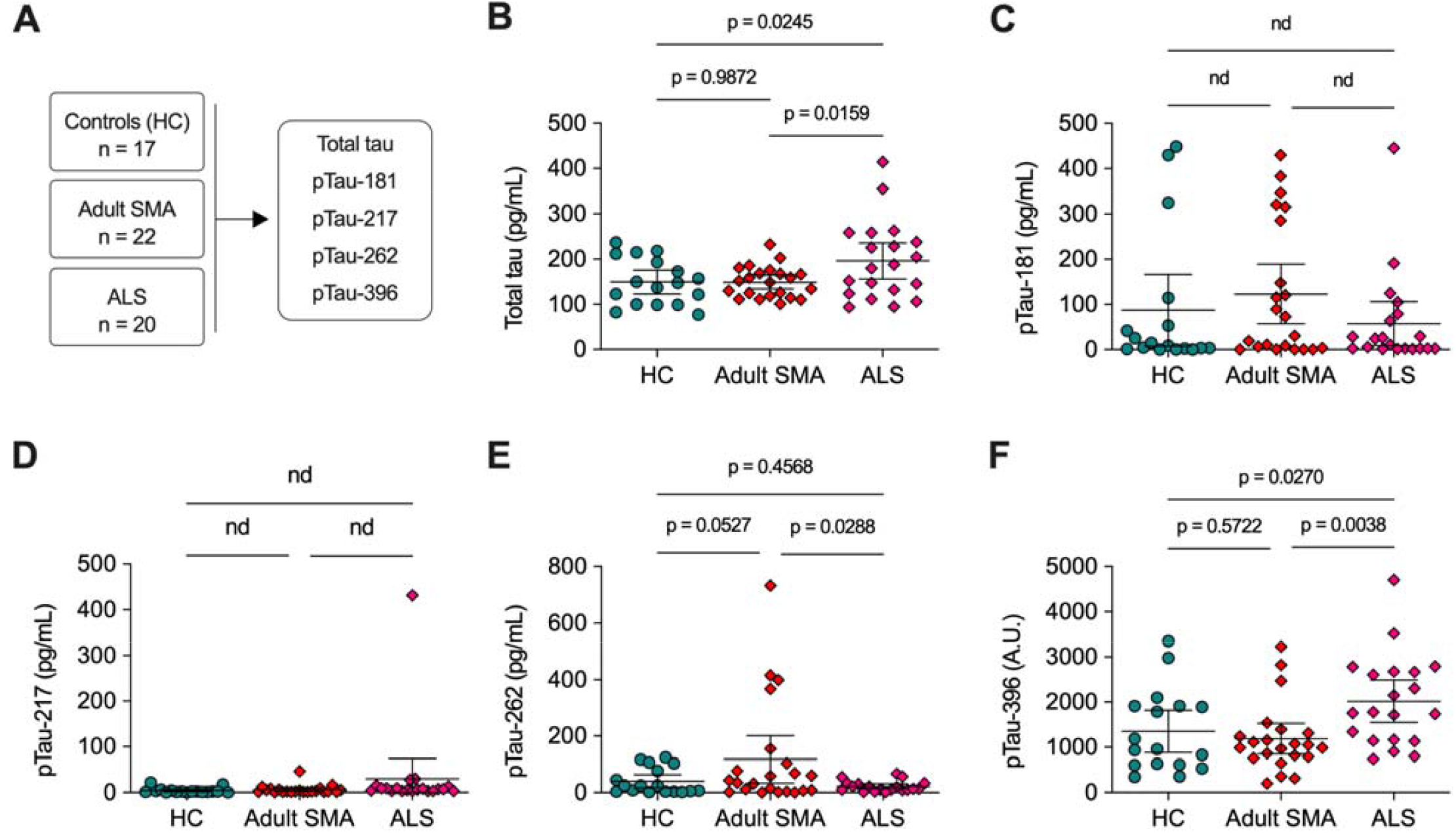
Circulating Tau levels in a cohort of adult subjects including healthy controls (n = 17), patients with SMA (n = 22), and patients with ALS (n = 20). **A**. Schematic overview of cohort composition and analytes measured from plasma samples. **B–F**. Concentrations of total tau (**B**), pTau-181 (**C**), pTau-217 (**D**), pTau-262 (**E**), and pTau-396 (**F**) across groups. Each point represents data from an individual participant; horizontal bars denote mean ± 95% confidence interval (CI). Statistical comparisons were performed using one-way ANOVA followed by multiple comparisons by controlling false discovery rate using two-stage step-up method of Benjamini, Krieger and Yekutieli. Critical value was set as p < 0.05. When one-way ANOVA demonstrated significant difference, p-values derived from multiple comparisons are shown. “nd” indicates no significant difference in one-way ANOVA or multiple comparisons.

We next examined circulating Tau measurements in a natural history cohort of pediatric SMA patients with 2 or ≥3 SMN2 copies and compared them with age-matched healthy controls (**Figure 2A**). Overall, no significant differences were observed for any Tau species assessed (**Figure 2A–F**). Notably, a small subset of the most severely affected infants with 2 SMN2 copies exhibited elevated pTau-181 (**Figure 2C**), pTau-262 (**Figure 2E**), and pTau-396 (**Figure 2F**) levels that were not seen in healthy controls, although these findings were not consistent across all SMA infants in this group. These data indicate that Tau alterations are not reliably detected in early life, even among the most severe SMA cases. However, the few exceptions observed underscore the value of closely monitoring each newly diagnosed patient.

**Figure 2.**
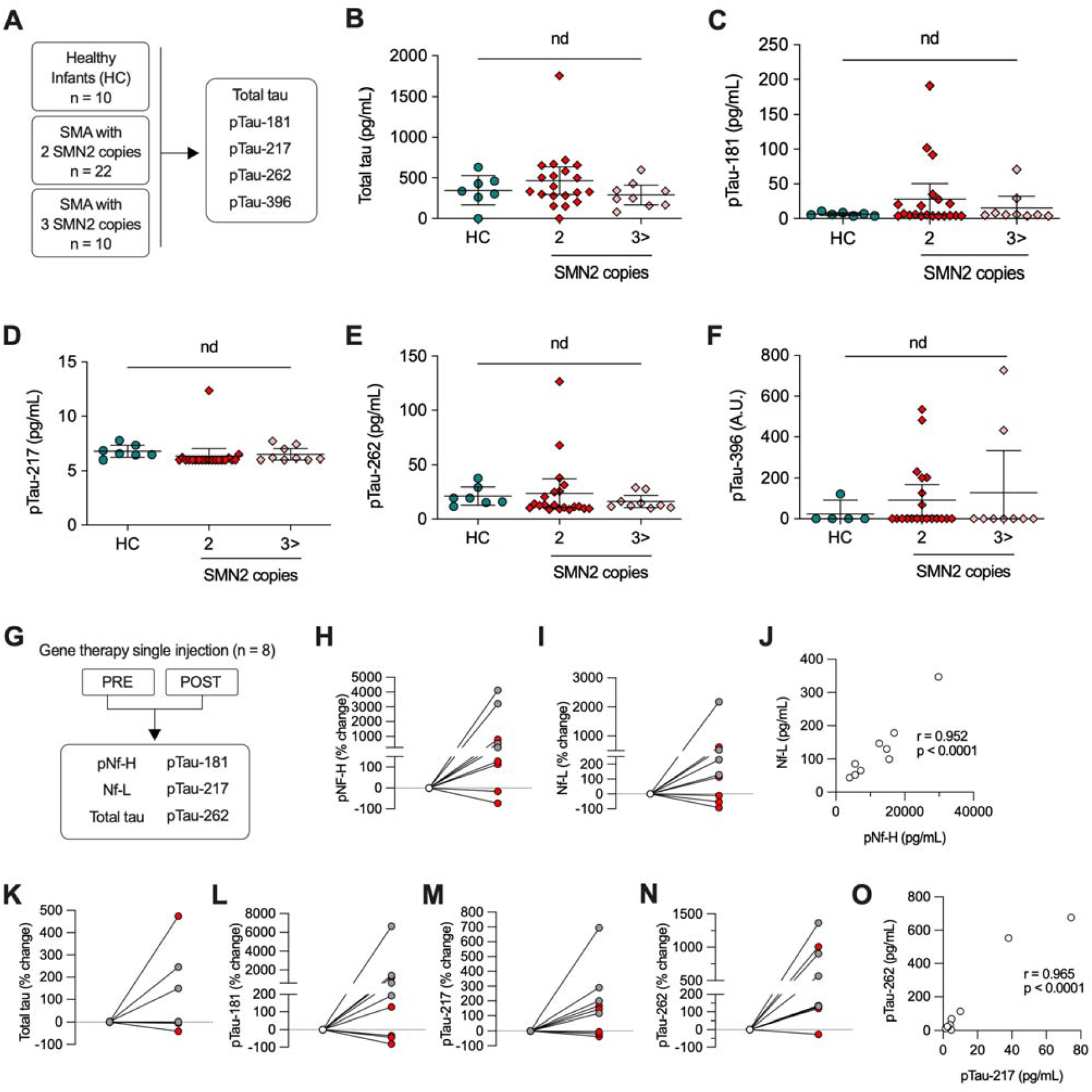
Circulating Tau levels in cohorts of pediatric subjects including healthy controls (n = 10), SMA infants with 2SMN2 copies (n = 22), and SMA infants with 3 or more SMN2 copies (n = 10). **A**. Schematic overview of cohort composition and analytes measured from plasma samples. **B–F**. Concentrations of total tau (**B**), pTau-181 (**C**), pTau-217 (**D**), pTau-262 (**E**), and pTau-396 (**F**) across groups. Each point represents data from an individual participant; horizontal bars denote mean ± 95% confidence interval (CI). Statistical comparisons were performed using one-way ANOVA followed by multiple comparisons by controlling false discovery rate using two-stage step-up method of Benjamini, Krieger and Yekutieli. Critical value was set as p < 0.05. When one-way ANOVA demonstrated significant difference, p-values derived from multiple comparisons are shown. “nd” indicates no significant difference in one-way ANOVA or multiple comparisons. **G**. Schematic of additional cohort including 8 infants with SMA treated with gene therapy (onasemnogene abeparvovec) early in life. Paired samples were obtained before (PRE) and up to 180 days after (POST) a single injection of onasemnogene abeparvovec. **H-O**. Percent change from baseline in pNF-H (**H**), Nf-L (**I**), total tau (**K**), pTau-181 (**L**), pTau-217 (**M**), and pTau-262 (**N**) following gene therapy only (gray) or co-therapy with nusinersen (red). Plots display individual trajectories, highlighting biomarker reductions or increases on an individual-subject level. **J** and **O**. Pearson correlations between pNf-H and Nf-L (**J**) or between pTau-217 and pTau-262 (**O**). Statistical significance and correlation coefficients are indicated on plots.

Given our previous observations that onasemnogene abeparvovec monotherapy is associated with increased neurofilament levels in symptomatic SMA infants early in life, we next sought to determine whether similar effects would be observed in circulating Tau measurements. We analyzed a subset of eight patients with available samples before treatment and up to six months following onasemnogene abeparvovec administration (**Figure 2G**). Among these, four patients received onasemnogene abeparvovec alone, while the remaining four were co-treated with nusinersen. Consistent with the neurofilament changes observed previously (**Figures 2H–J**), we detected increased Tau levels after onasemnogene abeparvovec infusion, with more pronounced elevations in patients receiving monotherapy compared with those on combination therapy (**Figure 2K-O**). These findings parallel our earlier unexpected neurofilament results and raise the hypothesis that gene therapy may induce acute collateral stress or damage in vulnerable neurons, potentially through enhanced cellular stress response^14^. Additional mechanistic studies will be required to clarify the biological basis of these observations.

## Discussion

We report real world data from a series of Tau measurements in different cohorts of SMA patients. Despite advances in newborn screening programs and availability of 4 FDA-approved therapies for SMA, there remains an urgent need for simple, sensitive, and specific biomarkers capable of tracking early disease progression and treatment response. In this context, our group and others have explored the use of circulating survival motor neuron (SMN) protein levels, electrophysiologic measures, circulating neurofilament levels among other candidates as complementary readouts to enhance clinical monitoring in SMA^13–15^. Despite important advances, all these candidates have specific limitations and additional efforts are necessary towards a more complete panel of biomarkers to track SMA progression^13–15^. Given its mechanistic role and the current findings, Tau represents a compelling candidate for further consideration in this context.

Our observations highlight several important considerations. First, pTau-262 emerged as a potentially SMA-specific alteration in adults, suggesting disease-relevant kinase dysregulation or selective susceptibility of spinal motor neurons. Second, the absence of consistent Tau alterations in untreated SMA infants likely reflects the distinct developmental physiology. Third, the transient Tau increases following gene therapy parallel neurofilament kinetics and raises the possibility of acute neuronal or synaptic vulnerability during vector transduction. Larger longitudinal cohorts, harmonized sample handling, and integration of Tau species with other molecular and physiologic markers is encouraged to define the utility of Tau as part of a multidimensional SMA biomarker framework.

## Data Availability

All data relevant to this study are contained within the manuscript.

## Acknowledgments

We are grateful to all the patients and families who participated in this study.

## Appendix 1.

Authors.

**Appendix 1. Authors.**
| **Name** | **Location** | **Role** | **Contribution** |
| --- | --- | --- | --- |
| Leillani L. Ha | Massachusetts General Hospital, Boston, MA | Author | Directed the research project and designed experiments; analyzed the data; wrote the manuscript; participated in the data interpretation, reviewed and approved the final manuscript. |
| Sunayana Mitra | Massachusetts General Hospital, Boston, MA | Author | Collected clinical data; analyzed the data; participated in the data interpretation, reviewed and approved the final manuscript. |
| Doreen T. Ho | Massachusetts General Hospital, Boston, MA | Author | Collected clinical data; participated in the data interpretation, reviewed and approved the final manuscript. |
| Becky Fillingham | Massachusetts General Hospital, Boston, MA | Author | Collected clinical data; participated in the data interpretation, reviewed and approved the final manuscript. |
| James D. Berry | Massachusetts General Hospital, Boston, MA | Author | Collected clinical data; participated in the data interpretation, reviewed and approved the final manuscript. |
| Kathryn J. Swoboda | Massachusetts General Hospital, Boston, MA | Author | Collected clinical data; participated in the data interpretation, reviewed and approved the final manuscript. |
| Christiano R. R. Alves | Massachusetts General Hospital, Boston, MA | Author | Directed the research project and designed experiments; collected clinical data; wrote the manuscript; participated in the data interpretation, reviewed and approved the final manuscript. |

## Author Contributions

L.L.H. and C.R.R.A. directed the research project. S.M., D.T.H., B.F., J.D.B. and K.J.S. collected clinical data. L.L.H., S.M., D.T.H., B.F., J.D.B., K.J.S., and C.R.R.A. analyzed the data or participated in the data interpretation. L.L.H. and C.R.R.A. drafted the manuscript. All authors reviewed and approved the final manuscript.

